# Individualized melanoma risk prediction using machine learning with electronic health records

**DOI:** 10.1101/2024.07.26.24311080

**Authors:** Guihong Wan, Sara Khattab, Katie Roster, Nga Nguyen, Boshen Yan, Hannah Rashdan, Hossein Estiri, Yevgeniy R. Semenov

## Abstract

**Background:** Melanoma is a lethal form of skin cancer with a high propensity for metastasizing, making early detection crucial. This study aims to develop a machine learning model using electronic health record data to identify patients at high risk of developing melanoma to prioritize them for dermatology screening.

**Methods:** This retrospective study included patients diagnosed with melanoma (cases), as well as matched patients without melanoma (controls), from Massachusetts General Hospital (MGH), Brigham and Women’s Hospital (BWH), Dana-Farber Cancer Institute (DFCI), and other hospital centers within the Research Patient Data Registry at Mass General Brigham healthcare system between 1992 and 2022. Patient demographics, family history, diagnoses, medications, procedures, laboratory tests, reasons for visits, and allergy data six months prior to the date of first melanoma diagnosis or date of censoring were extracted. A machine learning framework for health outcomes (MLHO) was utilized to build the model. Performance was evaluated using five-fold cross-validation of the MGH cohort (internal validation) and by using the MGH cohort for model training and the non-MGH cohort for independent testing (external validation). The Area Under the Receiver Operating Characteristic Curve (AUC-ROC) and the Area Under the Precision-Recall Curve (AUC-PR), along with 95% Confidence Intervals (CIs), were computed.

**Results:** This study identified 10,778 patients with melanoma and 10,778 matched patients without melanoma, including 8,944 from MGH and 1,834 from non-MGH hospitals in each cohort, both with an average follow-up duration of 9 years. In the internal and external validations, the model achieved AUC-ROC values of 0.826 (95% CI: 0.819–0.832) and 0.823 (95% CI: 0.809–0.837) and AUC-PR scores of 0.841 (95% CI: 0.834–0.848) and 0.822 (95% CI: 0.806–0.839), respectively. Important risk features included a family history of melanoma, a family history of skin cancer, and a prior diagnosis of benign neoplasm of skin. Conversely, medical examination without abnormal findings was identified as a protective feature.

**Conclusions:** Machine learning techniques and electronic health records can be effectively used to predict melanoma risk, potentially aiding in identifying high-risk patients and enabling individualized screening strategies for melanoma.

## INTRODUCTION

Despite significant therapeutic advancements in the treatment of late-stage melanoma, the projected number of melanoma-related deaths in the United States is expected to exceed 90,000 within the next decade.^1^ Early detection of melanoma is of paramount importance, as the five-year survival rate for patients with melanoma diagnosed at a localized stage is greater than 99% and falls to 35% when the disease metastasizes to distant organs.^2^ Routine dermatologic screening is one of the most effective ways to detect melanoma at early stages. The incidence rate of melanoma has been rising rapidly over the past few decades, with an estimated 100,640 cases of invasive melanoma and 99,700 cases of in situ melanoma to be diagnosed in the United States in 2024.^3^ These numbers are still significantly smaller than the entire population in the United States, which would make population-level screening too costly and impractical. Thus, there is a need for predictive models that enable healthcare providers to identify and enroll high-risk patients in melanoma screening programs.

The widespread adoption of electronic health record (EHR) system has led to the accumulation of an unprecedented amount of patient information, holding great potential for personalized medicine.^4^ However, utilizing EHR data with conventional analytic methods to build predictive tools has been challenging due to the large volume of data and the complexity of processes, which often contain irrelevant information.^5^ Recent advances in machine learning techniques have enabled feature mining,^6^ dimensionality reduction,^7^ and robust prediction of patient outcomes across many diseases.^8,9^ For example, a self-adaptive machine learning framework for health outcomes has been developed and successfully used to predict long-term sequelae of COVID-19.^9^ These techniques have not yet been applied to predict an individual’s risk of developing melanoma.

In this study, we aim to develop a machine learning model for identifying patients at high risk of melanoma development using EHR data from a large-scale multi-institutional registry. Since the data utilized in the model is collected from routine office visits, patients can be risk stratified without undergoing a specific assessment. This approach can be scaled with minimal cost to triage and identify high-risk patients for melanoma screening programs, enabling early disease detection and therapeutic intervention.

## METHODS

### Study Design

**Figure 1** presents the study concept. The outcome or event of interest in this study is the development of melanoma versus no melanoma. The respective event date is the date of first melanoma diagnosis for the melanoma group and date of death or last visit for the no melanoma group. The primary goal of the study is to predict the 6-month risk of melanoma development. In our secondary analyses, we conducted experiments using 3, 9, and 12 months as the time horizon for the prediction.

**Figure 1.**
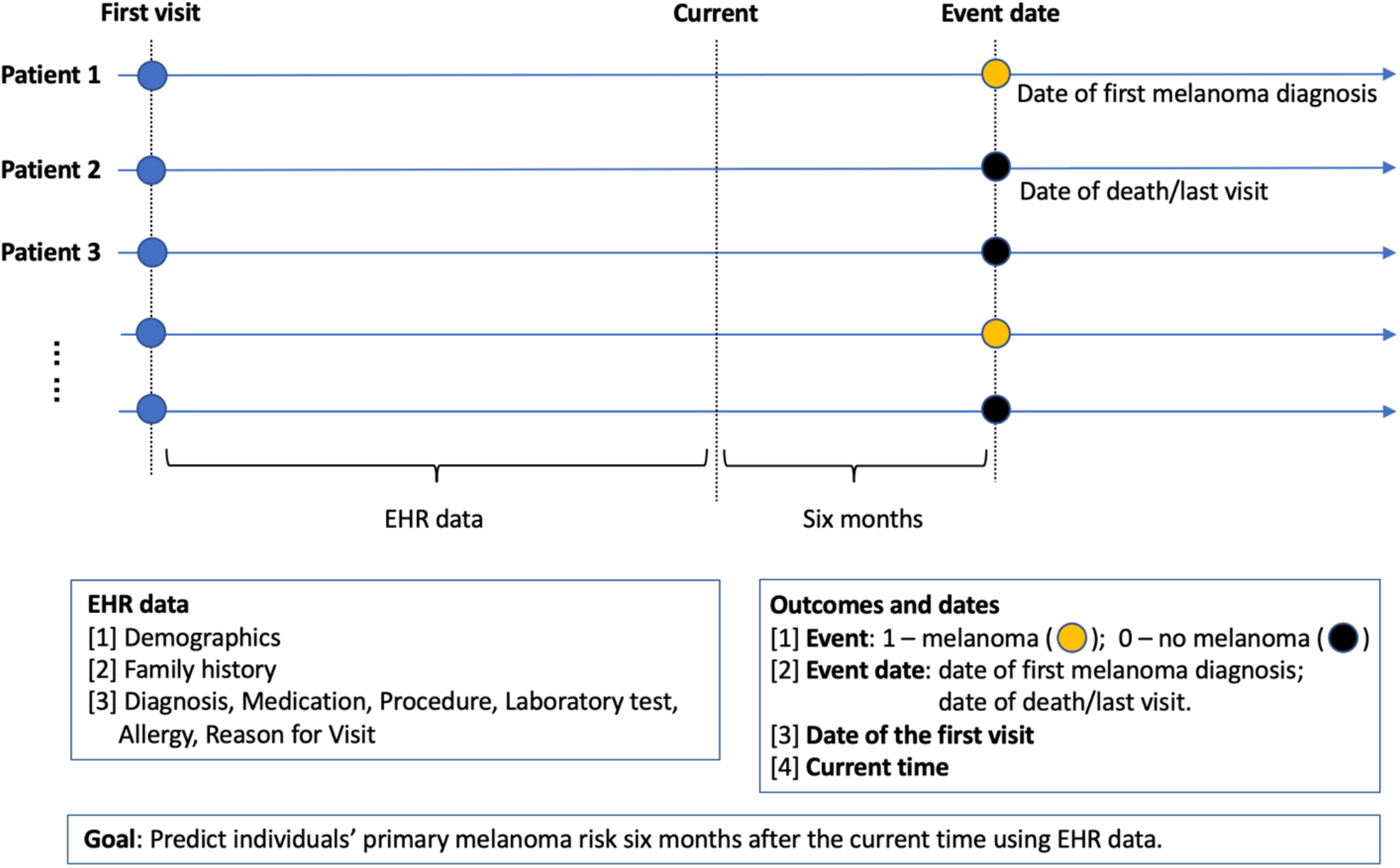
Study Design. The outcome or event of interest in this study is melanoma vs. no melanoma. Suppose the current time was six months before the event date, the objective of the study is to predict a patient’s melanoma risk using the patent’s Electronic Health Record (EHR) data before the current time.

### Patients and Data Collection

We leveraged the Research Patient Data Registry (RPDR), which is a clinical database at the Mass General Brigham healthcare system containing detailed data on over 12 million unique patients seen across the Massachusetts General Hospital (MGH), Brigham and Women’s Hospital (BWH), Dana-Farber Cancer Institute (DFCI), and other hospital centers. The aggregated data included patient demographics, reasons for visit, diagnoses, laboratory tests, and others. This retrospective study included patients diagnosed with melanoma between May 1992 and November 2022.

**Figure 2** presents the flow diagram illustrating the identification process of the study population. First, we identified all patients 18 years of age and older who were diagnosed with melanoma prior to November 15, 2022. A 1:3 matched cohort of non-melanoma patients was then identified based on age, sex, and race using the “match controls” function in the RPDR system. Due to the extremely large volume of patients in RPDR, it was not practical to include all non-melanoma patients. Following the application of exclusion criteria, the non-melanoma group was selected through 1:1 matching based on the duration from the first visit to the event time using the “MatchIt” R package (version 4.5.0).

**Figure 2.**
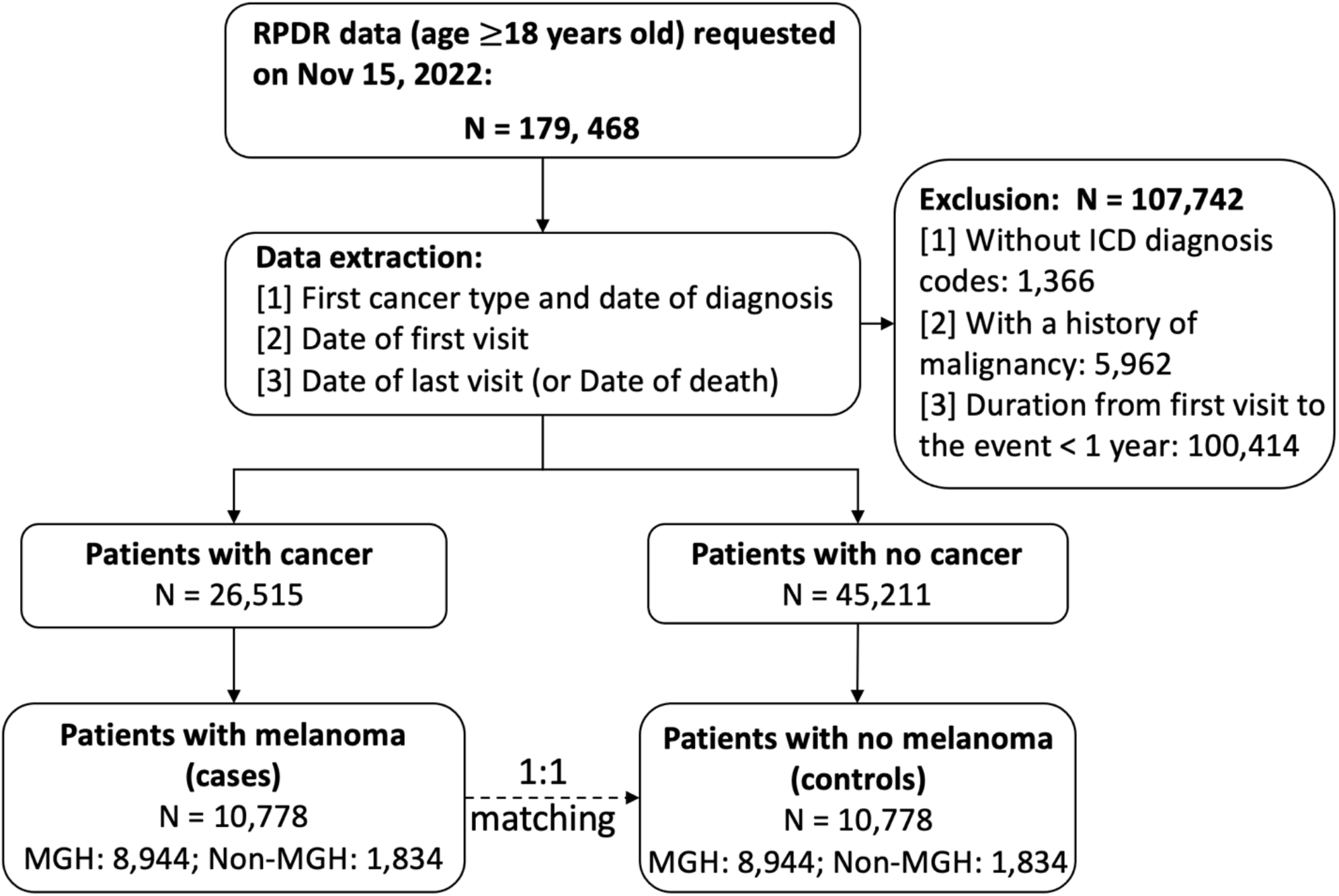
The study population. This retrospective study, started by requesting data from the Research Patient Data Registry (RPDR) at Mass General Brigham on November 15, 2022, including patients with melanoma diagnoses who were at least 18 years old. Additionally, a 1:3 matched cohort of non-melanoma patients, based on age, sex, and race, was included. (Due to the extremely large size of the RPDR, not all non-melanoma patients could be included.) Following the application of exclusion criteria, the study included 26,515 cancer patients and 45,211 non-cancer patients. Among them, 10,778 patients with melanoma were included, with an equal number of 10,778 patients without melanoma identified through 1:1 matching based on the duration from the first visit to the event time. Patients from Massachusetts General Hospital (MGH) were utilized for model development, while those from other hospitals (Non-MGH) served for external validation of the model.

We used diagnostic codes from the International Classification of Diseases, 9th Revision (ICD-9) and 10th Revision (ICD-10) to identify the first primary cancer and date. Specifically, patients with melanoma were identified by the ICD-9 code of 172 and the ICD-10 codes of C43 and D03. Patients who had secondary malignancies (ICD-9: 196-198; ICD-10: C77-C79 and C7B) or personal history of malignancies (ICD-9: V10; ICD-10: Z85) before the first primary cancer dates were excluded. To reduce the likelihood of false group labeling, patients with any cancer recorded by other non-ICD diagnostic codes (e.g., internal codes) were excluded.

We included the following data from the RPDR as features in the machine learning model: demographics (race, sex, age, marital status, US state), family history, diagnoses, medications, medical procedures, laboratory tests, reasons for visit, and allergy data. Patients without records of demographics and diagnoses in RPDR were excluded. To ensure sufficient features for modeling, patients with less than one year follow-up from first visit to the event were excluded. We also excluded evidently irrelevant codes, such as Encounter for Immunization (ICD-9: V04.81; ICD-10: Z23), and Established Patient Office Visit (CPT: 99213, CPT: 99214).

### Statistical Analyses

In this study, we leveraged the machine learning framework for health outcomes (MLHO) developed at Mass General Brigham.^9^ The conventional aggregated data (e.g., count of individual diagnoses) of the training set were fed into the framework, followed by dimensionality reduction and feature selection. The resulting features were used to build a binary classification model. We compared the performance of models built with three machine learning algorithms: eXtreme Gradient Boosting (xgbTree; in the xgboost package, version 1.5.0.2), gradient boosting machines (gbm; in the gbm package, version 2.1.8), and generalized linear model (glm; in the stats package, version 3.6.3). We evaluated models in two ways: (1) five-fold cross-validation on the MGH cohort (referred to as internal validation); (2) using the MGH cohort for model training and validating the model independently with patients from other hospitals (Non-MGH, mainly BWH/DFCI). Two threshold-free metrices were used to measure a model’s performance: Area Under the Receiver Operating Characteristic Curve (AUC-ROC) and Area Under the Precision-Recall Curve (AUC-PR), along with 95% Confidence Intervals (CIs), were computed.

Furthermore, SHapley Additive exPlanations (SHAP) values were used to investigate how much each feature contributes to model predictions.^10^ Features with positive SHAP values positively impact the prediction, while those with negative values have a negative impact. SHAP values are zero for missing or irrelevant features. Features were ranked based on average absolute SHAP values across all patients in the training set.

To compare the characteristics of two groups, we conducted Pearson’s Chi-squared tests for categorical variables and Student’s t-tests for continuous variables. All statistical analyses were conducted using R statistical software (version 3.6.3).

## RESULTS

### Participant Characteristics

This study identified 10,778 patients with melanoma and 10,778 matched patients without melanoma, including 8,944 from MGH and 1,834 from non-MGH hospitals in each cohort, both with an average follow-up duration of 9 years (**Supplementary Table 1**). There were more females (54.4% vs. 51.1%; p<0.001) and White patients (92.5% vs. 81.4%; p<0.001) in the melanoma group compared to the no melanoma group. Patients in the melanoma group were younger (57 vs. 61 years old; p<0.001) than patients in the no melanoma group.

**Table 1** shows the machine learning model performance in the 6-month early prediction. The model built using xgbTree achieved best performance (AUC-ROC: 0.826, 95% CI: 0.819– 0.832; 0.823, 95% CI: 0.809–0.837 and AUC-PR: 0.841, 95% CI: 0.834–0.848; 0.822, 95% CI: 0.806–0.839) in the internal and external validations, respectively. The model performances and the receiver operating characteristic curves for 3-month, 6-month, 9-month, and 12-month early detections are presented in **Figure 3**. There were no significant differences among the different month early detections (p>0.05). Important risk features included family history of melanoma, benign neoplasm of skin, and family history of skin cancer. Conversely, medical examination without abnormal findings was identified as a protective feature **(Figure 4**).

**Table 1.** Machine learning model performance in the six-month early prediction.

| Algorithm | Metrics | Training | Internal | External |
| --- | --- | --- | --- | --- |
| **xgbTree** | AUC-ROC<br>[95% CI] | 0.864<br>[0.859-0.869] | 0.826<br>[0.819-0.832] | 0.823<br>[0.809-0.837] |
|  | AUC-PR<br>[95% CI] | 0.876<br>[0.871-0.882] | 0.841<br>[0.834-0.848] | 0.822<br>[0.806-0.839] |
| **gbm** | AUC-ROC<br>[95% CI] | 0.834<br>[0.828-0.839] | 0.822<br>[0.816-0.829] | 0.816<br>[0.802-0.829] |
|  | AUC-PR<br>[95% CI] | 0.847<br>[0.840-0.854] | 0.835<br>[0.828-0.843] | 0.815<br>[0.798-8.834] |
| **glm** | AUC-ROC<br>[95% CI] | 0.812<br>[0.806-0.819] | 0.802<br>[0.796-0.809] | 0.797<br>[0.783-0.812] |
|  | AUC-PR<br>[95% CI] | 0.826<br>[0.819-0.834] | 0.817<br>[0.809-0.825] | 0.799<br>[0.782, 0.818] |

Training: performance on the training (MGH) cohort; Internal: five-fold cross validation on the MGH cohort; External: training on the MGH cohort and validation on the Non-MGH cohort. AUC-ROC: Area under the Receiver Operating Characteristic Curve; AUC-PR: Area under the Precision-Recall Curve; CI: Confidence Interval.

**Figure 3.**
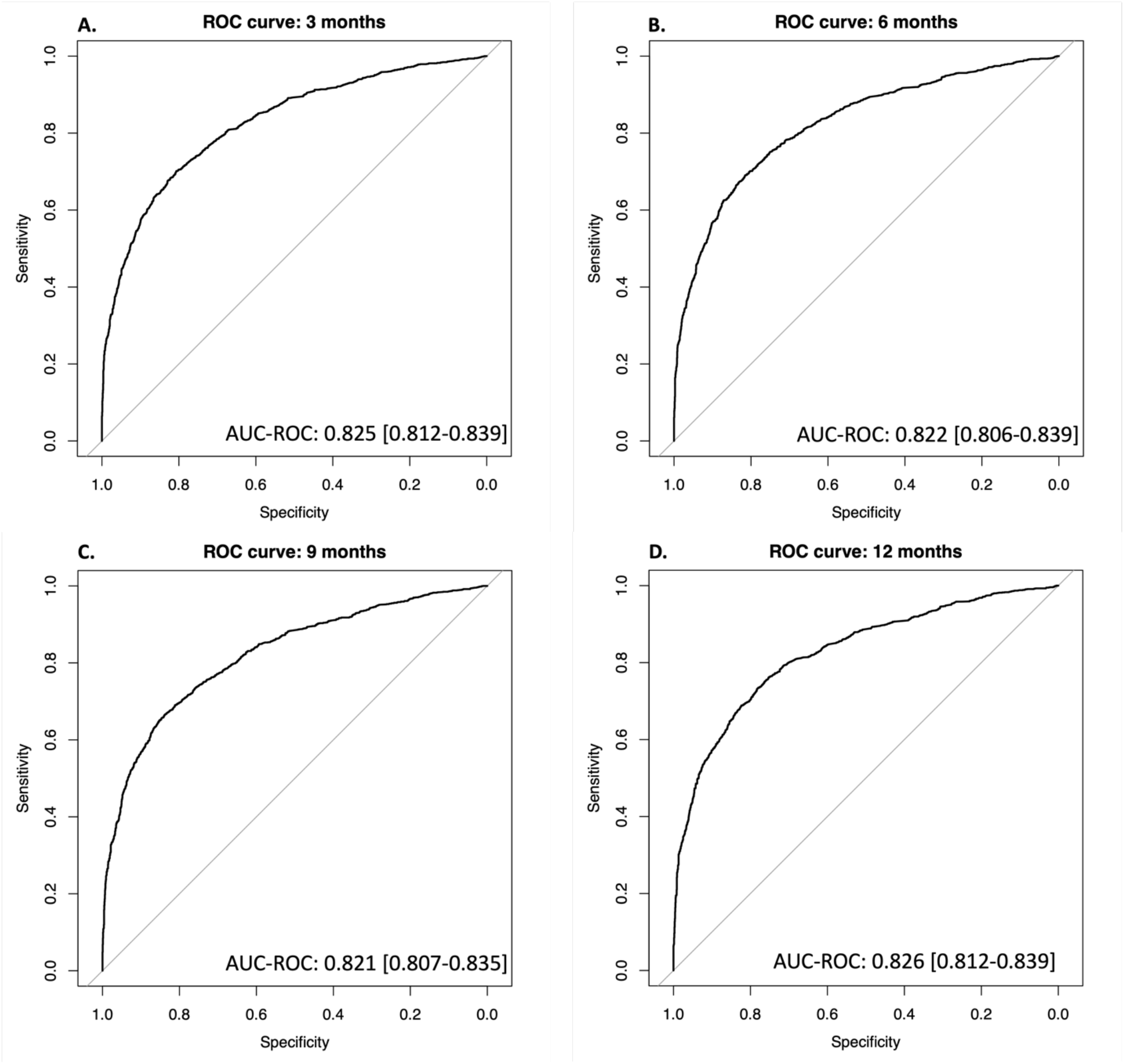
The ROC curves of the xgbTree models for predicting melanoma risk. **A**. The ROC curve of three-month early prediction in the external validation. **B**. The ROC curve of six-month early prediction in the external validation. **C**. The ROC curve of nine-month early prediction in the external validation. **D**. The ROC curve of twelve-month early prediction in the external validation. ROC: receiver operating characteristic.

**Figure 4.**
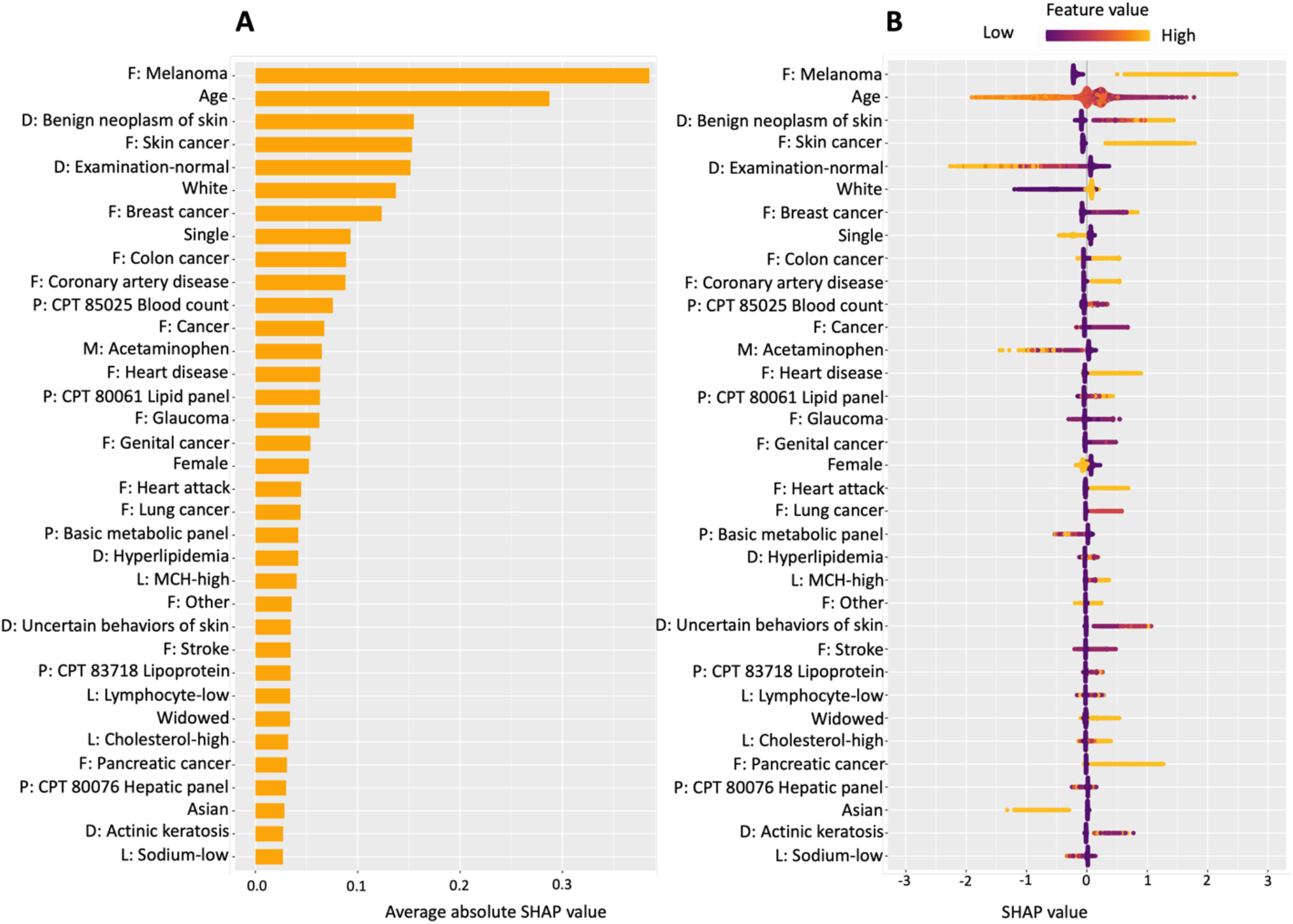
Top 35 important features in the xgbTree model in the six-month early prediction. Positive SHAP value indicates an increased risk of melanoma. The family history of melanoma (F: Melanoma) was identified as the most significant risk factor. Patients whose medical examination without abnormal findings (D: Examination-normal) had a decreased risk of melanoma. “F:” represents Family History; “D:” represents Diagnosis; and “L:” represents Laboratory Test. “P:” represents Procedure. MCH: Mean Corpuscular Hemoglobin.

**Supplementary Table 2** presents the model performances with different EHR modules. The baseline (using demographics alone) AUC-ROC value was 0.643 (95% CI: 0.625–0.661) in the external validation. When combining with family history, the model performance was significantly improved (AUC-ROC: 0.749, 95% CI: 0.727–9.771, p<0.001), while there was no improvement when adding the allergy data (AUC-ROC: 0.637, 95% CI: 0.600–0.674, p>0.05). **Supplementary Figure 1** presents the top 10 features in each EHR module.

## DISCUSSION

The increasing incidence of melanoma underscores the critical need for advanced methodologies utilizing risk prediction tools to identify high-risk patients and prioritize them in screening. In this study, we leveraged EHR data from a multi-institutional registry to evaluate the effectiveness of machine learning in predicting melanoma risk and identifying the most influential predictive features. Utilizing our extensive and longitudinal dataset, our xgbTree model demonstrated the most robust performance in both internal and external validations.

Our findings demonstrate that machine learning models have the potential to reliably identify individuals at heightened risk for melanoma using EHR data, as has been shown in studies of other primary malignancies such as in lung and breast cancer.^11,12^ Previous research utilizing machine learning techniques to forecast susceptibility to cancer have reported AUC values ranging from 0.648 in breast cancer to 0.89 in non-melanoma skin cancer risk predication models, which demonstrates the robustness of this model in relation to previously published findings.^12,13^ Our investigation identified family history of melanoma as the predominant risk factor in assessing an individual’s susceptibility to melanoma, followed by a past diagnosis of benign neoplasms of the skin, a family history of skin cancer, and White race. While established risk factors such as familial history and race are well-recognized in melanoma risk assessment, our comprehensive dataset and machine learning model unveiled several additional risk factors that were previously unknown or inadequately established. Notably, family history of breast cancer and colon cancer were identified among the most important predictive factors in a patient’s medical history. We suspect that these risk factors are due to familial genetic cancer syndromes, such as BRCA2, which have been postulated to increase risk of melanoma development.^14^ Additionally, having had a prior medical examination without abnormal findings was identified as a protective feature, which may be attributed to primary care providers often being the first to raise concerns regarding lesions that appear irregular.

In this study, we present the computational foundation for an EHR-based triage tool which could have significant clinical applicability in guiding personalized screening strategies for primary melanoma development. This tool is valuable for clinical practice as it allows for the identification of patients at high risk of developing melanoma without needing to screen the entire population, which is also neither recommended according to the recent United States Preventive Services Task Force statement nor feasible.^15,16^

Limitations of this study include its retrospective nature, resulting in some variables being unavailable for certain patients. Additionally, our models were developed using patients from a geographically similar area. It should also be noted that the melanoma and the non-melanoma groups were 1:1 matched to balance the cohorts for model training and facilitate interpretation of model accuracy. However, the real-world incidence of melanoma is much smaller. Future studies should utilize a more diverse cohort, incorporate measures of unstructured features (e.g., prior ultraviolet light exposure), and consider explicitly incorporating germline susceptibilities to melanoma to strengthen the discriminatory power of the model and enhance accuracy of screening recommendations.

## Data Availability

All summary data supporting the findings of this study available within the article or its supplementary materials. The patient data generated for this study can only be shared per specific institutional review board requirements. Upon request to the corresponding author, a data sharing agreement can be initiated following institution-specific guidelines.

---

**Supplementary Table 1.**
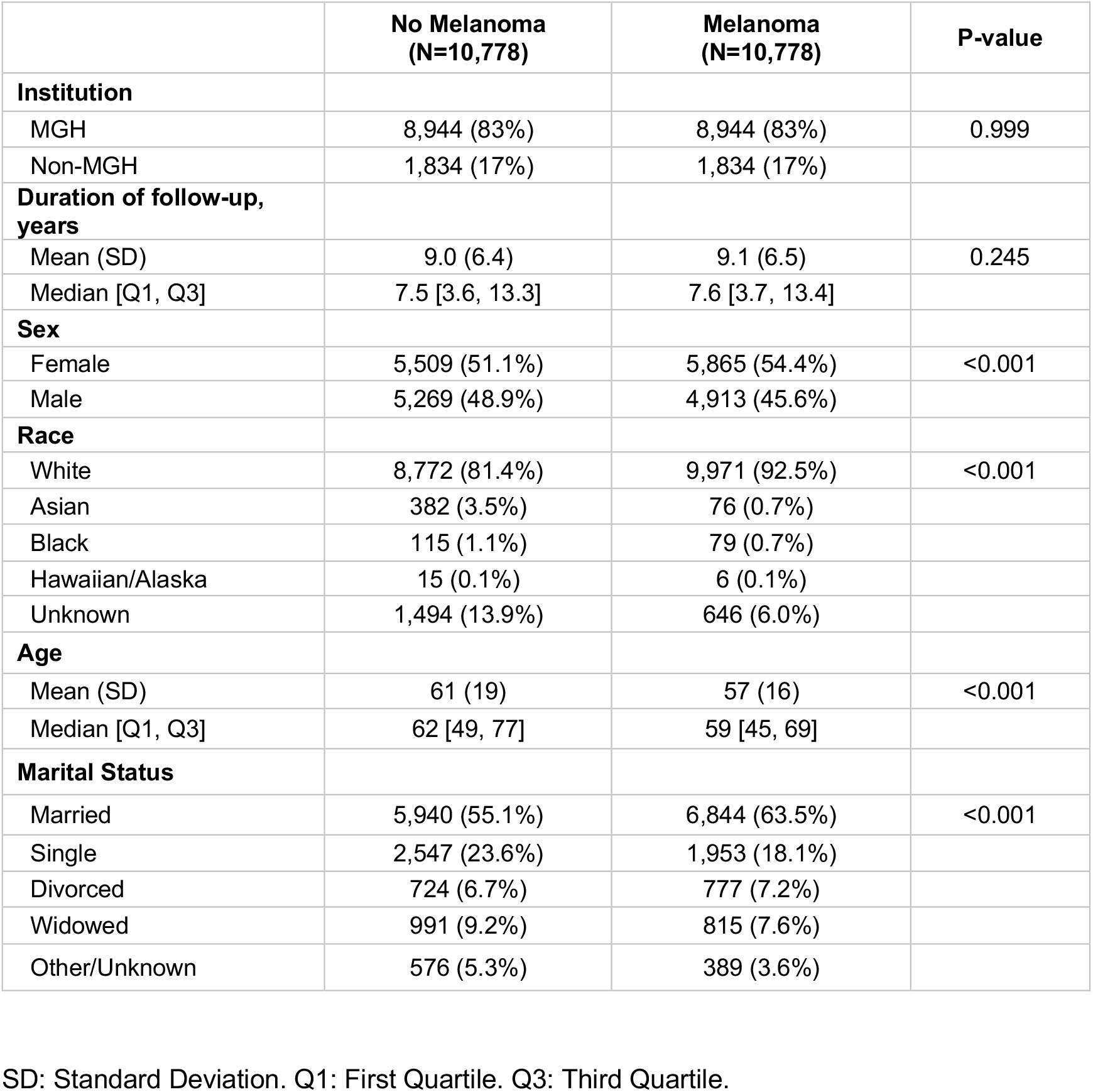
Basic characteristics of the study population.

**Supplementary Table 2.** Model performance with different EHR data.

|  | **Demographics**<br>**(Baseline)** | **Demographics +**<br>**Family History** | **Demographics +**<br>**Diagnoses** | **Demographics +**<br>**Procedures** |
| --- | --- | --- | --- | --- |
| **Training** | 0.686<br>[0.678-0.693] | 0.819<br>[0.811-0.828] | 0.791<br>[ 0.785-0.798] | 0.792<br>[0.785-0.798] |
| **Internal** | 0.671<br>[0.663-0.679] | 0.795<br>[0.786-0.804] | 0.741<br>[0.734-0.749] | 0.736<br>[0.728-0.744] |
| **External** | 0.643<br>[0.625-0.661] | 0.749<br>[0.727-0.771] | 0.729<br>[ 0.711-0.748] | 0.724<br>[0.704-0.743] |
|  | **Demographics +**<br>**Laboratory Tests** | **Demographics +**<br>**Medications** | **Demographics +**<br>**Allergies** | **Demographics +**<br>**Reasons for**<br>**Visit** |
| **Training** | 0.773<br>[0.765-0.781] | 0.789<br>[0.781-0.798] | 0.725<br>[0.709-0.741] | 0.766<br>[0.743-0.789] |
| **Internal** | 0.729<br>[0.720-0.738] | 0.749<br>[0.739-0.758] | 0.710<br>[0.694-0.726] | 0.726<br>[0.701-0.750] |
| **External** | 0.692<br>[0.671-0.714] | 0.684<br>[0.661-0.707] | 0.637<br>[0.600-0.674] | 0.706<br>[0.647-0.765] |

The result format is AUC-ROC [95% CI]. EHR: electronic health record. AUC-ROC: area under the receiver operating characteristic curve. CI: confidence interval.

**Supplementary Figure 1.**
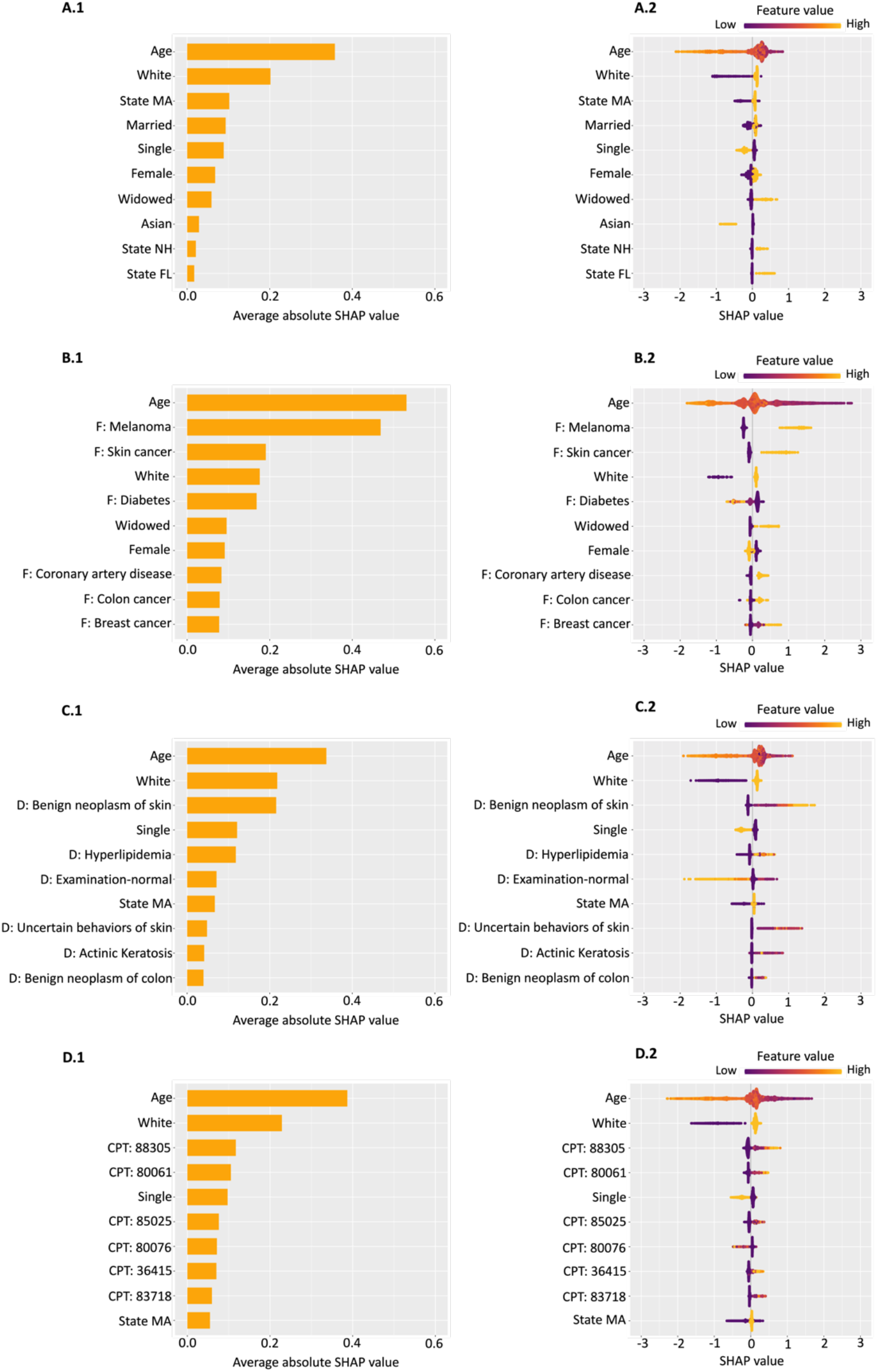
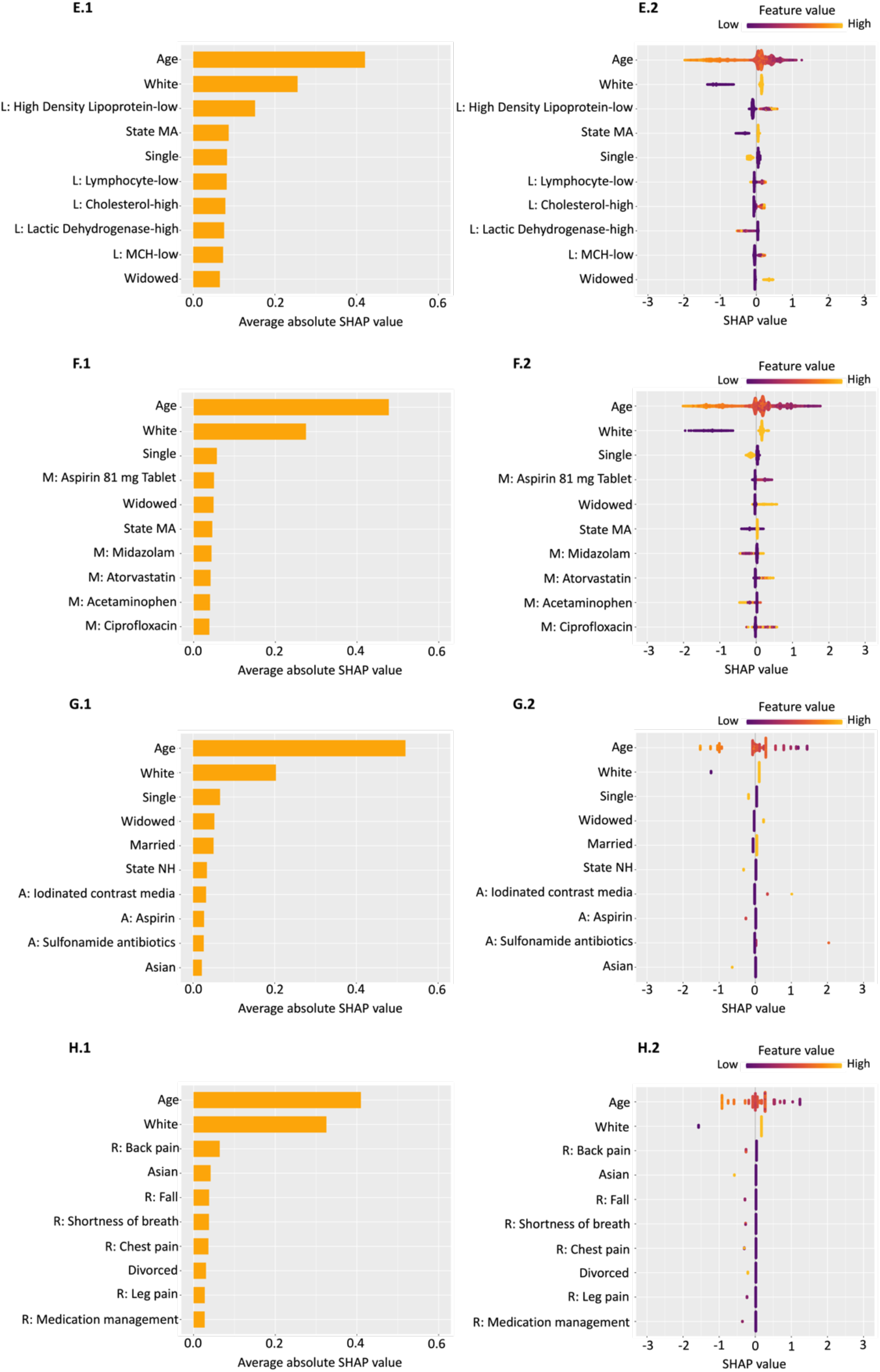
Top 10 important features in the six-month early prediction. CPT 88305: Surgical pathology, gross and microscopic examination. CPT 80061: Lipid panel. CPT 85025: Blood count. CPT 80076: Hepatic function panel. CPT 36415: Collection of venous blood by venipuncture. CPT 83718: Lipoprotein, direct measurement. **A.1** and **A.2**: Demographics. **B.1** and **B.2**: Demographics + Family History. **C.1** and **C.2**: Demographics + Diagnoses. **D.1** and **D.2**: Demographics + Procedures. **E.1** and **E.2**: Demographics + Laboratory Tests. **F.1** and **F.2**: Demographics + Medications. **G.1** and **G.2**: Demographics + Allergies. **H.1** and **H.2**: Demographics + Reasons for Visit.

